# Oral Chinese medicines for treating diabetic macular edema: Protocol for a systematic search of randomized studies and meta-analysis

**DOI:** 10.1101/2023.04.04.23288114

**Authors:** Miaoran Gao, Sheng Huang, Jian Zhou, Yiqiu Yang, Xianke Luo, Changlu Yang, Xinning Yu, Mengdan Tang, Xiaoling Yan

## Abstract

**Introduction:** The diabetic macular edema (DME) is a relevant cause of visual impairment in diabetes. The current treatments are limited by high costs, risk of infections and damage to retinal cells. Randomized controlled studies (RCTs) have investigated oral traditional Chinese medicines (TCMs) for the treatment of DME. We aimed at determining the efficacy and safety of oral TCMs by systematically reviewing the full set of studies.

**Methods and analysis:** Published RCTs will be searched through 12 databases until October 1, 2022. Two investigators will conduct independent literature search, data extraction and assessment of quality. The risk of bias will be judged with the version 2 of the Cochrane risk-of-bias tool. The RevMan software will be utilized to analyze data. Dichotomous data will be assessed by using odds ratios and 95% confidence intervals (CIs). We will evaluate continuous data by using weighted mean differences and 95% CIs. We are going to assess heterogeneity by Cochran’s Q test and the I^2^ statistics. We plan sensitivity analysis and subgroup analysis to identify sources of heterogeneity. Funnel plots, Egger’s tests and Begg’s tests will be also performed.

**Protocol registration number:** The study protocol is registered on PROSPERO (CRD42022379268).

**Strengths and limitations:**

- This is the first systematic study examining the efficacy and safety of orally administered Chinese medicines for DME.
- We will search randomized controlled trials in 12 databases.
- We will implement subgroup analysis and sensitivity analysis to explore any source of heterogeneity.
- The presence of several types of macular oedema might challenge the workload for this study.

## Introduction

Diabetic macular edema (DME) affects approximately 1 out of 15 adults with diabetes [1,2]. In accordance with the International Diabetes Federation, the number of adults living with diabetes is estimated to reach 783 million by 2045 [3]. The high incidence of DME seriously affects quality of life and puts a financial burden on healthcare systems.

Although the pathogenesis is unclear, the current understanding of DME focuses on inflammation, vascular lesions and neurodegeneration [4]. The treatments consist of anti-vascular endothelial growth factor (VEGF) therapies, corticosteroids and laser therapy. Prior of the implementation of anti-VEGF medicines, macular laser was the mainstay of treatment. However, laser photocoagulation has been associated with irreversible retinal damages [5]. Intravitreal anti-VEGF agents are nowadays prescribed as first-line treatment in several countries [6-9]. The clinical response is variable, with the incidence of refractory DME reaching up to 50% of treated individuals [10]. Frequent intravitreal injections of anti-VEGF medicines are often required to keep long-term vision, increasing the risk of endophthalmitis and healthcare expenditures [11]. The switching from anti-VEGF medications to corticosteroids often improves the therapeutic outcome in the long-term [12,13], but intravitreal steroids can cause high intraocular pressure and cataract [14-16].

The treatment of DME requires affordable, safe and effective therapeutic options. Previously, oral traditional Chinese medicines (TCM) have been described as potential therapies against DME [17,18]. However, randomized controlled trials (RCTs) displayed inconsistent results [19-21]. This study aimed at systematically evaluating the benefits of oral TCM for DME. Our study will support the establishment of effective, safe and personalized treatment strategies for DME.

## Materials and methods

### Study protocol

The study protocol is registered on PROSPERO (CRD42022379268). The protocol complies with the Cochrane Manual [22] and follows the Preferred Reporting Items for Systematic Review and Meta-Analysis Protocols (PRISMA-P) guidelines [23,24].

### Patient and public involvement

No patient took part in the study design, conception and conduction of this research.

### Search strategy

Independently, two investigators (MG and SH) will search for studies in the China National Knowledge Infrastructure (CNKI), the Wanfang Database, the China Biomedical Literature Service System (SinoMed), the Chinese Scientific and Technical Journals Database (VIP), Web of Science, PubMed, OVID, EMBASE, Scopus, MedRxiv, PsyArXiv and BioRxiv. The search will end on October 1, 2023 with no language restrictions. In addition, ongoing and unpublished trials will be searched in the International Clinical Trials Registry Platform (ICTR). Studies included in the OpenGrey database will be explored using manual searches. The following MeSH terms and keywords will be employed: herbal medicine, macular, edema, macular edema, diabetic macular edema, diabetes, type 2 diabetes, type 1 diabetes, traditional Chinese medicine, Chinese herbal medicine, clinical randomized controlled trials, randomized, randomized controlled trials, controlled clinical trials, pharmacotherapy, and placebo. The search will be first conducted on PubMed with the search strategy displayed in Table 1. Then, the search will be redesigned to the different databases. Any inconsistency will be solved with the opinion of a third investigator (XY).

**Table 1.** Search strategy for PubMed

| NO. | Search term |
| --- | --- |
| #1 | [MeSH Terms]Macular edema |
| #2 | Macular odema |
| #3 | Edema,macular |
| #4 | Odema,macular |
| #5 | Diabetic macular edema |
| #6 | Diabetic macular odema |
| #7 | Macular edema,diabetic |
| #8 | Macular odema,diabetic |
| #9 | DME |
| #10 | DMO |
| #11 | Macular edema in type 1 diabetes |
| #12 | Macular odema in type 1 diabetes |
| #13 | Macular edema in type 2 diabetes |
| #14 | Macular odema in type 2 diabetes |
| #15 | Macular edema , type 1 diabetes |
| #16 | Macular odema , type 1 diabetes |
| #17 | Macular edema , type 2 diabetes |
| #18 | Macular odema , type 2 diabetes |
| #19 | #1-#18/OR |
| #20 | [MeSH Terms] Medicine, Chinese traditional |
| #21 | Chinese Medicine, Traditional |
| #22 | Chinese Traditional Medicine |
| #23 | Traditional Chinese Medicine |
| #24 | Traditional Medicine, Chinese |
| #25 | Drugs, Chinese Herbal |
| #26 | Chinese Drugs, Plant |
| #27 | Chinese Herbal Drugs |
| #28 | Herbal Drugs, Chinese |
| #29 | Herbal Medicine |
| #30 | Medicinal Herbs |
| #31 | Herb, Medicinal |
| #32 | Medicinal Herb |
| #33 | Herbs, Medicinal |
| #34 | Medicine, Herbal |
| #35 | Herbal Medicines |
| #36 | #20-#35/OR |
| #37 | groups [Title/Abstract] |
| #38 | trial [Title/Abstract] |
| #39 | randomized controlled trial [Publication Type] |
| #40 | controlled clinical trial [Publication Type] |
| #41 | RCT |
| #42 | randomized [Title/Abstract] |
| #43 | randomly [Title/Abstract] |
| #44 | placebo [Title/Abstract] |
| #45 | drug therapy [MeSH Subheading] |
| #46 | #37-#45/OR |
| #47 | animals [MeSH Terms] NOT humans [MeSH Terms] |
| #48 | #46 NOT #47 |
| #49 | #19 AND #36 AND #46 AND #48 |

### Eligibility criteria

#### Study design

This systematic review will include RCTs of any duration and sample size irrespective of language, year of publication and publication status.

#### Participants

We will include participants diagnosed with DME and treated with oral TCMs. No restrictions on age, gender or ethnicity are planned. Patients with different types of macular edema will be excluded. We will not include trials without clear diagnostic criteria.

#### Interventions

Oral Chinese medicines will be considered experimental interventions, with no restrictions on the type of Chinese medicine. Placebo, reference drugs (e.g., anti-VEGF or steroids) or usual care will be considered control interventions. If oral Chinese medicines are combined with other treatments in the experimental group, the control group should display similar treatments.

#### Outcomes

The primary outcomes are as follows: best corrected visual acuity (BCVA), central macular concave thickness (CMT) and total effective rate (number of patients healed and/or in remission in each group divided by the number of patients). The secondary outcomes are: incidence rate of adverse events (e.g., high intraocular pressure or intraocular inflammation), incidence rate of macular edema recurrence and change in planimetric area of fluorescein leakage from retinal neovascularization using fundus fluorescein angiography.

#### Exclusion criteria

The exclusion criteria are: studies with invalid or missing data, studies with non-standardized outcome evaluation, non-randomized controlled trials, preclinical studies, case reports or reviews, republished studies (we will retain the most comprehensive one) and studies of low quality (risk of bias score below 3).

### Study selection and data extraction

#### Study selection

MG and SH will independently import the articles in the Endnote X9 software. Titles, abstracts and full texts will be screened once duplicates are removed. Disputes will be solved with the opinion of a third researcher (XY). The PRISMA flowchart illustrates the process of study selection (Figure 1).

**Figure 1.**
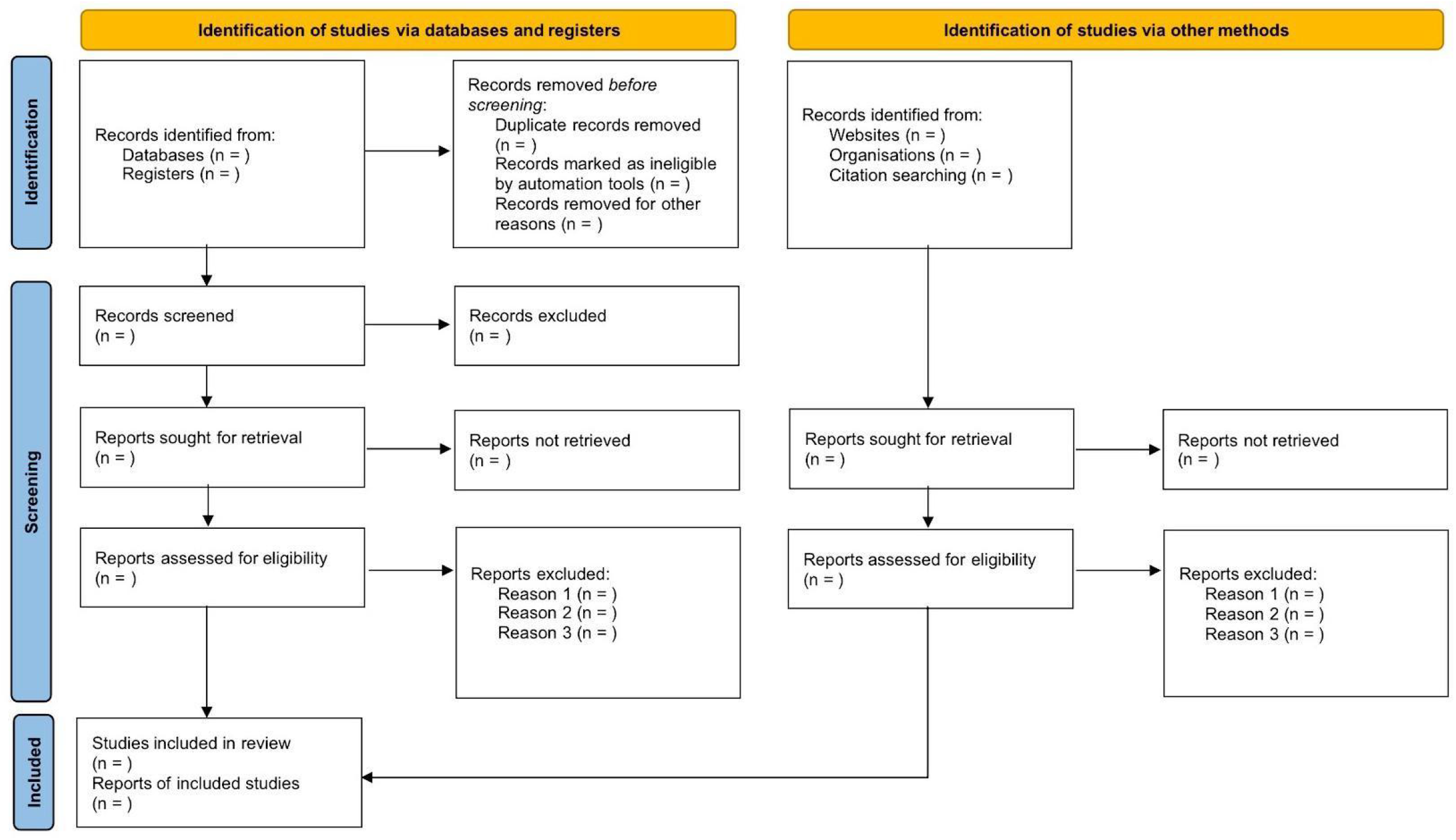
PRISMA flflow diagram for identifying, screening, and determining the eligibility and inclusion of studies.

#### Data extraction

Two investigators (MG and SH) will independently extract information using a standardized, pilot-tested form. The extracted data include (1) general information, such as title, author, sample size and year of publication; (2) patient information, including gender and age; (3) study interventions and (4) outcome indicators, including CMT, BCVA, total effective rate, adverse events, recurrence rate. Discrepancies will be discussed with a third researcher (XY). Final data will be integrated in the Review Manager.

#### Quality assessment

MG and SH will assess the risk of bias of each trial utilizing the version 2 of the Cochrane risk-of-bias tool (RoB 2.0). The evaluation includes methods of random allocation, allocation concealment, blinding of study participants and investigators, blinding of study assessors, missing outcomes, selective reporting and other potential bias. Judgments of “high risk”, “unclear risk” or “low risk” will be made for these seven aspects. Any disagreement will be discussed with a third researcher (XY).

### Data synthesis and statistical analysis

#### Data synthesis

We are going to use the RevMan 5.4.1 software for meta-analysis data interpretation. Mean difference (MD) and relative risk (RR) will be expressed with 95% confidence intervals (95% CIs). The Cochrane Handbook will be used for guidance. A *p* value < 0.05 will be deemed statistically significant.

#### Heterogeneity

Heterogeneity will be explored by using the χ^2^ test (test level α = 0.1), while the magnitude will be determined quantitatively with I^2^. We will utilize a fixed effects model if no significant heterogeneity is reported between studies. Otherwise, we will utilize a random effects model.

#### Subgroup analysis

We will explore possible sources of heterogeneity based on: (1) trial information, such as first author, sample size, study design and year of publication; (2) patient information, including gender, age, nationality and disease duration; (3) type of oral TCM, formulation (decoction or granules) and single or combined treatments (such as an oral TCM combined with an anti-VEGF); (4) type of intervention implemented in the control group and (5) treatment duration.

#### Sensitivity analysis

We are going to perform a sensitivity analysis through a one-by-one exclusion method assessing the impact of each individual study on the effect size.

#### Publication bias

We will appraise publication bias through the Egger’s and Begg’s test. The funnel chart method will be used to visualize the bias. In addition, we will use the trim-and-fill method to correct the funnel asymmetry.

#### Quality of evidence

We will utilize the GRADE criteria (high, medium, low and very low) to assess the quality. The quality for outcome indicators will be appraised primarily in terms of risk of study bias, publication bias, inconsistency, indirectness and imprecision [25]. Two researchers (MG and SH) will independently score each domain and discrepancies will be solved by discussion.

## Results

The results will be presented by using narrative summaries, flowcharts, summary tables and statistical analyses (with meta-analysis when possible).

## Discussion

DME is a significant cause of vision impairment in diabetes [26,27]. The occurrence of DME involves multiple pathological processes and different signaling pathways. VEGF is a mediator causing macular edema and retinal neovascularization [28-30]. Specifically, VEGF phosphorylates vascular endothelial cadherin (VE-cadherin) plus upregulates nitric oxide (NO) expression. In DME, chronic hyperglycemic states and pro-inflammatory cytokines lead to a pathological increase of VEGF [29,31,32]. Chronic hyperglycemia causes retinal inflammation by activating microglial cells, which produce pro-inflammatory cytokines and chemokines. Pro-inflammatory cytokines and other inflammatory mediators favor leukocyte adhesion and release of free radicals, enzymes and other substances that directly damage vascular endothelial cells [33]. Chemokines stimulate the secretion of cytokines exacerbating retinal vascular leakage, including VEGF, angiotensin II, interleukins and tumor necrosis factor-α [34]. Compared to healthy individuals, the levels of VEGFR-1, interleukin-6, interleukin-8, intercellular cell adhesion molecule-1 and recombinant macrophage inflammatory protein 1 beta were higher in patients with DME [35].

In Chinese medicine, DME is described as the “thirsty eye disease”, a disorder characterized by blood stasis and Qi deficiency [36]. Oral TCMs inhibit inflammation, improve microcirculation, reduce angiogenesis, enhance immune activities and regulate glucose metabolism [37-39]. For example, isoflavones in Pueraria lobata inhibit aldose reductase activity, reduce sorbitol pathway activity and lower blood glucose levels [40-42]. Astragalus inhibits the expression of several proteins related to the MAPK pathway and the secretion of pro-inflammatory molecules by reducing glial cell activation [43]. Gui Zhi regulates the expression of inflammatory mediators through Toll-like receptors and the nuclear factor-κB (NF-kB) pathway [44]. Atractylodes inhibit glial cell activity, reducing the expression of inflammatory factors and improving mitochondrial homeostasis [45]. Mimosa pudica is a plant associated with anti-inflammatory, immunomodulatory, hypoglycemic and antioxidant properties [46]. Poria inhibit their expression level, namely TNF-α, NF-KB, PI3K, AKT and other inflammation-related factors [47]. Salvia miltiorrhiza showed potential therapeutic effects on various models of inflammatory disorders by inhibiting TNF-α, NF-kB and other molecular targets [48]. Chaihu exerts a powerful anti-inflammatory activity by impairing the NF-kB pathway [49].

Several RCTs demonstrated benefits of oral TCMs in the prevention and treatment of DME [50-55]. Due to the absence of systematic studies evaluating oral TCMs in patients with DME, this study will provide a comprehensive evaluation of the available literature. This study will support clinicians to make informed decisions in patients with DME.

## Conclusions

Evidence on treating DME with oral TCMs is needed to assist physicians and other healthcare professionals in making informed clinical decisions. This study will potentially identify a new therapeutic option preventing and treating DME.

## Supporting information

PRISMA-P-checklist and Certificate of Funding

## Data Availability

All data produced in the present work are contained in the manuscript

## Ethics and dissemination

This meta-analysis involves primary literature, thus the ethical approval is not necessary. The final results will be reported in PROSPERO and peer-reviewed journals. The results will be shared through academic conferences if needed.

## Author Contributions

Conceptualisation: Miaoran Gao, Sheng Huang.

Data curation: Miaoran Gao, Sheng Huang, Jian Zhou, Yiqiu Yang, Xianke Luo, Changlu Yang, Xinning Yu, Mengdan Tang, Xiaoling Yan.

Formal analysis: Miaoran Gao, Sheng Huang, Jian Zhou, Yiqiu Yang. Funding acquisition: Jian Zhou, Xiaoling Yan.

Investigation: Yiqiu Yang, Xianke Luo, Changlu Yang. Methodology: Jian Zhou, Yiqiu Yang.

Supervision: Miaoran Gao, Sheng Huang, Xiaoling Yan. Validation: Yiqiu Yang, Xianke Luo, Mengdan Tang.

Writing the first draft: Miaoran Gao, Sheng Huang.

Writing, reviewing and editing: Changlu Yang, Xinning Yu, Xiaoling Yan.

## Funding

This work is funded by the National Natural Science Foundation of China (grant No. 81874491).

The funders will not have a role in study design, collection of data, data analysis and preparation of the article.

## Competing interests

No competing interests are reported by investigators.

## Patient consent for publication

Not applicable.

## Provenance and peer review

This study was not commissioned and will be externally peer reviewed.

## Notes

### Competing Interest Statement

The authors have declared no competing interest.

### Author Declarations

This is a clinical research design protocol

