## Supplementary material for "Oral Chinese medicines for treating diabetic macular edema: Protocol for a systematic search of randomized studies and meta-analysis": PRISMA-P-checklist and Certificate of Funding: Certificate of Funding.docx

**Notice on the approval of projects supported by the national natural science foundation of China**

Zhou Jian	Mr./Ms. ：

According to the Regulations of NSFC and the comments of experts, NSFC (hereinafter referred to as NSFC) decides to approve your application for funding. Project Approval Number: 81874491，Project Name: The effect of "Qingmang No.1 Prescription" on Retinal Ganglion Cells after Experimental Optic Nerve Injury ， Direct costs: 550 thousand yuan , Project Start/End Date: January 2019 to December 2022 ,The review comments and modification comments of relevant projects are attached.

National Natural Science Foundation of China

Medical Sciences

August 16, 2018
